# Variation in SARS-CoV-2 bioaerosol production in exhaled breath

**DOI:** 10.1101/2021.07.20.21260892

**Authors:** Renu Verma, Eugene Kim, Nicholas Degner, Katharine S. Walter, Upinder Singh, Jason R. Andrews

## Abstract

Using face mask bioaerosol sampling, we found substantial variation between individuals in SARS-CoV-2 copies exhaled over a 15-minute period, which moderately correlated with nasal swab viral load. Talking was associated with a median of 2 log_10_ greater exhaled viral copies. Exposure varies substantially between individuals but may be risk stratified by nasal swab viral load and whether the exposure involved conversation.

## Background

The Centers for Disease Control and Prevention (CDC) guidelines define a close contact as an individual who spent at least 15 minutes, over a 24-hour period, within 2 meters of an individual with COVID-19 (1, 2). This definition has been widely used to inform contact investigations in community as well as in healthcare settings (3) based on a premise of average risk, though the empirical basis for this specific time window is limited. We hypothesized that individual characteristics and actions- such as talking may also affect the overall bioaerosol shedding and might impact exposure risk over this time window.

Potential variation in SARS-CoV-2 bioaerosol production has been modeled by combining estimates of concentrations in airway swabs with data on total droplet production volume, but direct measurements of variation in exhaled SARS-CoV-2 abundance have been lacking (4). This may be in part due to lack of a flexible bioaerosol sampling tool which allows quantitative assessment of the determinants of bioaerosol variation within individuals. Face mask sampling is a convenient, low-cost bioaerosol sampling method which has been proven effective in detecting viruses including influenza and SARS-CoV-2 (5-7). However, these preliminary studies reported marginal sensitivities (38% to 40%) and additional data are required on the sensitivity of mask sampling. Additionally, mask sampling has not been explored as a tool to quantitatively assess the determinants of viral shedding. With improved sensitivity and high sample recovery, mask sampling can be used as an alternate sample source to study infectiousness. In this study, we developed a mask sampling tool to quantify and sequence SARS-CoV-2 from exhaled breath and used this to investigate the impact of speech and individual characteristics on viral shedding.

## The study

We recruited COVID-19 positive individuals from Stanford Hospital, including inpatient wards and an outpatient clinical trials unit, between September 2020 and March 2021 (**Table 1**). All participants were >18 years of age and provided informed consent. The study was approved by the Stanford IRB (#57686). We fitted N95 masks with a 47mm Petri dish (Fisherbrand) containing gelatine membrane filter (Sartorius) (**Supplementary figure 1**). We collected 141 mask samples from 97 individuals recruited in two groups (A & B). Group A (n=53 wore a mask for 30 minutes and was allowed to talk (or not) without further prompting. Group B (n=44) was instructed to wear two masks for 15 minutes each. For the first mask, they were instructed not to talk, while for the second mask they were asked to talk with the interviewer or a family member.

**Table 1:**
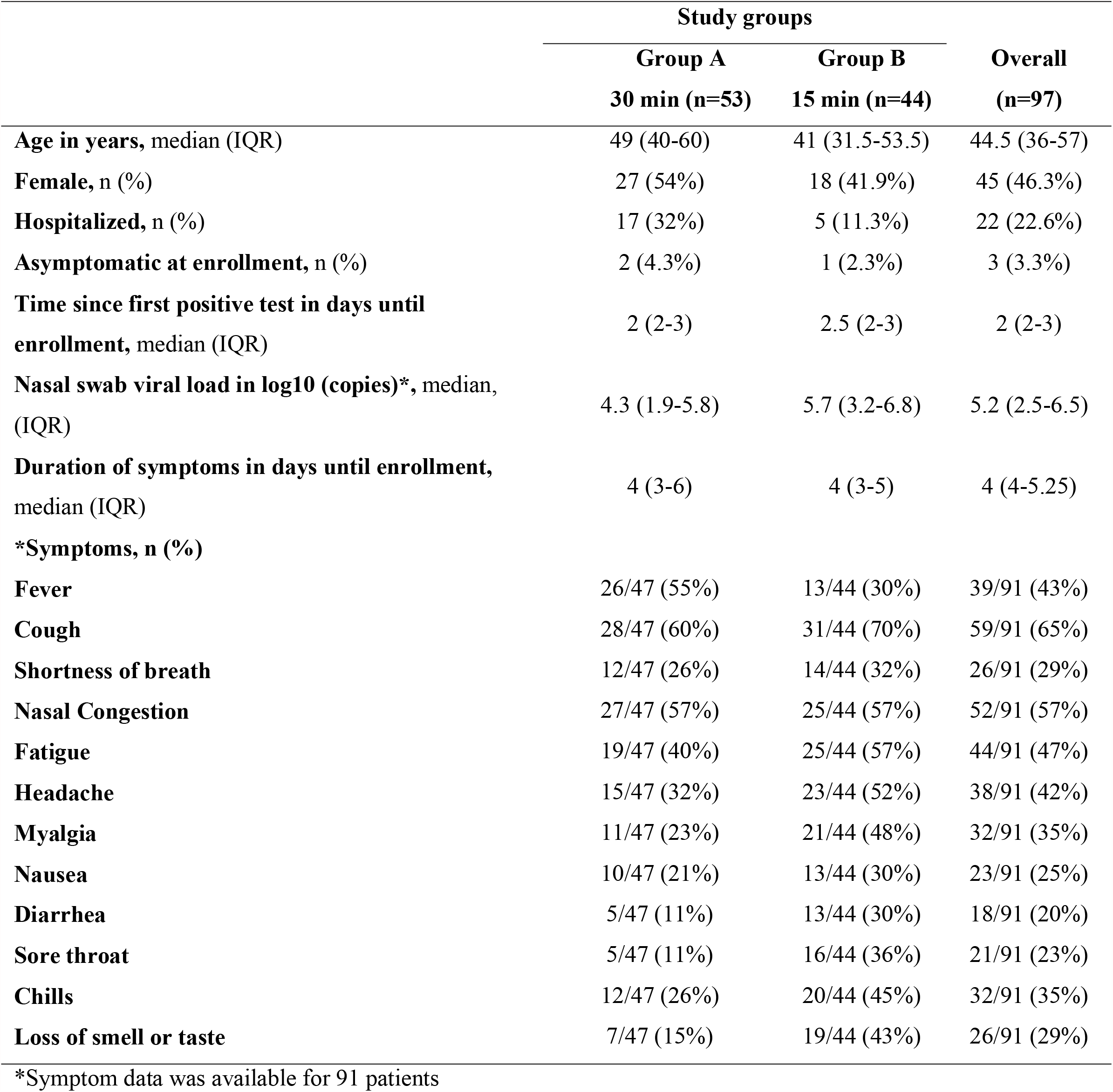
Study population characteristics.

|  | Study groups |  | Overall<br>(n=97) |
| --- | --- | --- | --- |
|  | Group A<br>30 min (n=53) | Group B<br>15 min (n=44) |  |
| **Age in years, median (IQR)** | 49 (40-60) | 41 (31.5-53.5) | 44.5 (36-57) |
| **Female, n (%)** | 27 (54%) | 18 (41.9%) | 45 (46.3%) |
| **Hospitalized, n (%)** | 17 (32%) | 5 (11.3%) | 22 (22.6%) |
| **Asymptomatic at enrollment, n (%)** | 2 (4.3%) | 1 (2.3%) | 3 (3.3%) |
| **Time since first positive test in days until enrollment, median (IQR)** | 2 (2-3) | 2.5 (2-3) | 2 (2-3) |
| **Nasal swab viral load in log10 (copies)\*, median, (IQR)** | 4.3 (1.9-5.8) | 5.7 (3.2-6.8) | 5.2 (2.5-6.5) |
| **Duration of symptoms in days until enrollment, median (IQR)** | 4 (3-6) | 4 (3-5) | 4 (4-5.25) |
| **\*Symptoms, n (%)** |  |  |  |
| **Fever** | 26/47 (55%) | 13/44 (30%) | 39/91 (43%) |
| **Cough** | 28/47 (60%) | 31/44 (70%) | 59/91 (65%) |
| **Shortness of breath** | 12/47 (26%) | 14/44 (32%) | 26/91 (29%) |
| **Nasal Congestion** | 27/47 (57%) | 25/44 (57%) | 52/91 (57%) |
| **Fatigue** | 19/47 (40%) | 25/44 (57%) | 44/91 (47%) |
| **Headache** | 15/47 (32%) | 23/44 (52%) | 38/91 (42%) |
| **Myalgia** | 11/47 (23%) | 21/44 (48%) | 32/91 (35%) |
| **Nausea** | 10/47 (21%) | 13/44 (30%) | 23/91 (25%) |
| **Diarrhea** | 5/47 (11%) | 13/44 (30%) | 18/91 (20%) |
| **Sore throat** | 5/47 (11%) | 16/44 (36%) | 21/91 (23%) |
| **Chills** | 12/47 (26%) | 20/44 (45%) | 32/91 (35%) |
| **Loss of smell or taste** | 7/47 (15%) | 19/44 (43%) | 26/91 (29%) |

\*Symptom data was available for 91 patients

The filter was dissolved in 1ml of Primestore MTM (Longhorn diagnostics) media and RNA was extracted using MagMAX™ Ultra Kit (Applied Biosystems). SARS-CoV-2 was detected with RT-qPCR using the CDC qualified N-gene assays and human RNaseP was used as a quality control (8). RNA recovery and limit of detection (LoD) from mask and swab were compared by spiking serially diluted SARS-CoV-2 synthetic RNA. SARS-CoV-2 genome was sequenced in-house from 21 exhaled breath and 14 paired nasal swab samples using ARTIC v3 Illumina sequencing protocol described previously (9) (**Supplementary methods**). In paired masks from the same individual, we compared SARS-CoV-2 detection dichotomously using McNemar’s test and quantitatively using Wilcoxon sign rank tests with continuity correction. We used the nfcore/viralrecon bioinformatic pipeline containerized on Nextflow to perform variant calling and generate consensus sequences from raw reads (10). We used the R package ape to measure pairwise SNP distance between consensus sequences (11).

SARS-CoV-2 was detected in exhaled breath from 71% (69/97) of participants, who were sampled at a median of 4 days (IQR: 3-5.25) from symptom onset and 2 days (IQR: 2-3) from first positive SARS-CoV-2 test. Among 77 patients with a nasal swab collected at the same encounter, 67 (87%) had SARS-CoV-2 positive nasal swabs. Mask samples were positive in 74.6% (50/67) of participants with positive swabs and 1/10 (10%) participants with negative swabs. Viral copy numbers in masks collected for 30 minutes (Pearson’s r = 0.76, p<0.001) and for 15 minutes (Pearson’s r = 0.58, p<0.001) talking were moderately correlated with paired nasal swabs (**Figure 1 A**). In group B (n=44), compared with mask samples collected while participants were not talking, mask samples collected while talking were more likely to be positive (59.0% vs 42.1%; p=0.061) and viral copies were significantly higher (median log_10_ difference: 2, IQR: 0, 3.6; p<0.001). Over 15 minutes of collection, total viral copies captured by masks varied substantially, ranging from 0 to 5.0 × 10^6^ copies (median= 393) while talking to 0 to 8.4 × 10^4^ copies (median= 0) while not talking **(Figure 1B)** (**Table 2**).

**Table 2:** SARS-CoV-2 N gene copies and RNaseP cycle threshold (Ct) values detected in paired samples from 44 individuals while talking or not talking for 15 minutes.

|  | Talking | No talking | P-value |
| --- | --- | --- | --- |
| **N-gene copies/mask\***, median (range) | 393 (0-5.1 x10<sup>6</sup>) | 0 (0-8.4x10<sup>4</sup>) | 0.028 |
| **RNaseP Ct**, median for Ct (range) | 32.2 (24.9-ND) | 34.6 (22.2-ND) | <0.001 |

\*Averaged copy numbers for N1 and N2 assays
†Wilcoxon signed rank test with continuity correction used to test significance
ND: Not detected

**Figure 1.**
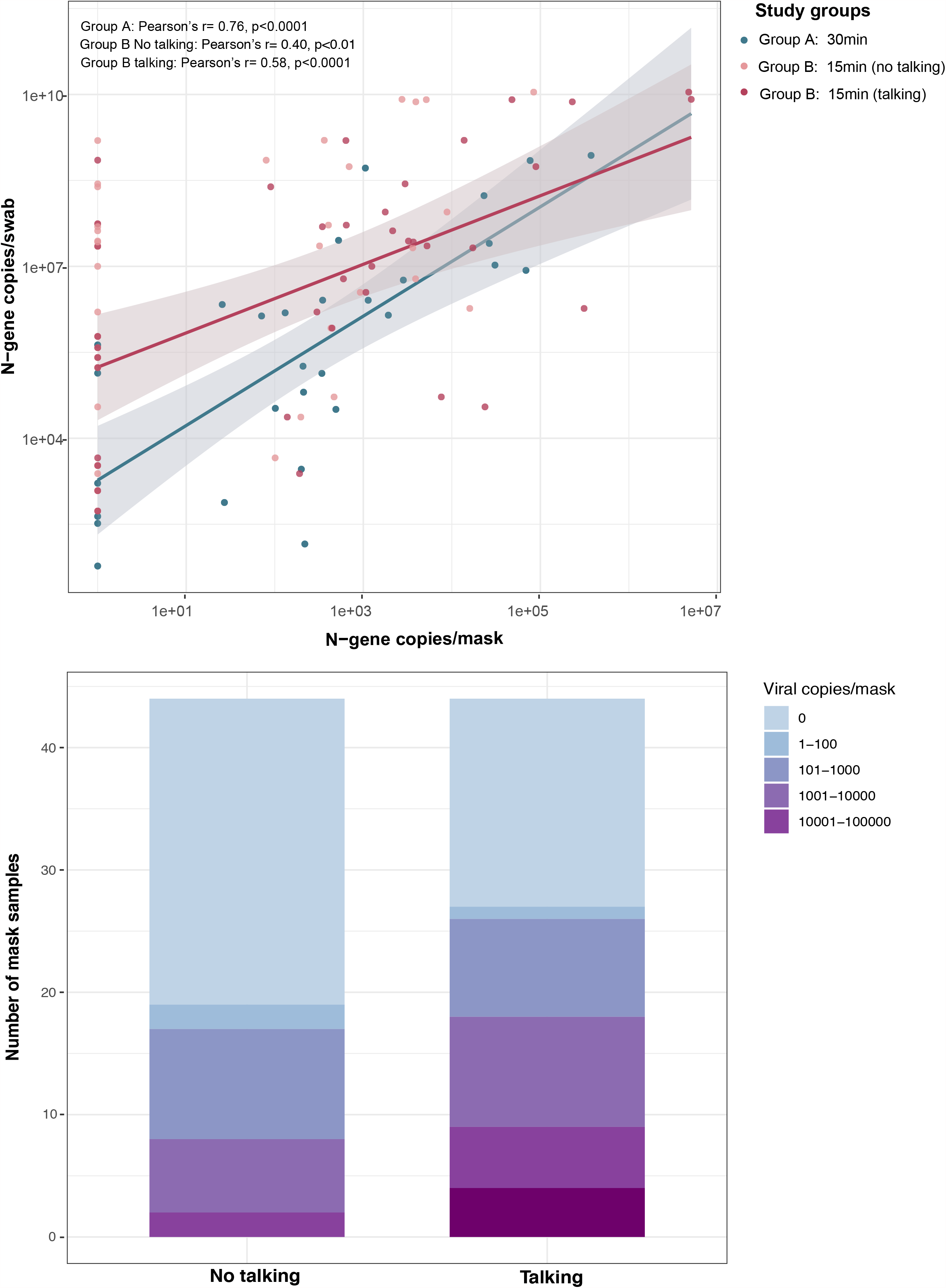
Correlation between mask and nasal swab viral copies. **(A)** Viral copy numbers detected in masks and nasal swabs were moderately correlated in group A (30 minutes sampling without instructions; blue, n=33) and group B (15 minutes sampling; red, n=44) talking cohort. **(B)** Total number of viral copies detected per mask sample in talking and no talking group. The viral copies were detected using SARS-CoV-2 specific N1 and N2 probes. Viral copies were quantified on standard curve derived from N-gene containing pET21b+ plasmid.

We further investigated the effect of age, sex and presence of symptoms with the number of viral particles released in exhaled breath. Viral load (Pearson’s r=0.57; p=0.001) and RNaseP abundance (p=0.04) in exhaled breath were moderately correlated with greater age in the 15- minute talking group. Overall, duration of symptoms (p=0.625), days from first positive SARS-CoV-2 test (p=0.812), and Human RNAseP copies (p=0.227) were not predictive of viral copies collected in masks. Mask positivity in inpatients 11/22 (50%) was significantly lower compared with outpatients 58/75 (77 %) (p=0.04).

To assess the quality of RNA extracted from the mask filters and explore the potential for mask sampling to be used for genomic surveillance, we sequenced the exhaled breath samples. The majority 71% (15/21) of all mask samples met our targets for sequencing coverage depth (100X) and width (>90% of the genome with >10X coverage) targets (**Figure 2A**). Median coverage depth was correlated with viral load, (Pearson’s r = 0.65, p<0.001) (**Figure 2B**). Phylogenetic (Pango) lineages assigned with sequences from mask samples were concordant with lineages assigned with sequences from nasopharyngeal swabs in all (14/14) paired samples meeting our coverage thresholds, including reported variants of concern (**Supplementary Table 1**).

**Figure 2.**
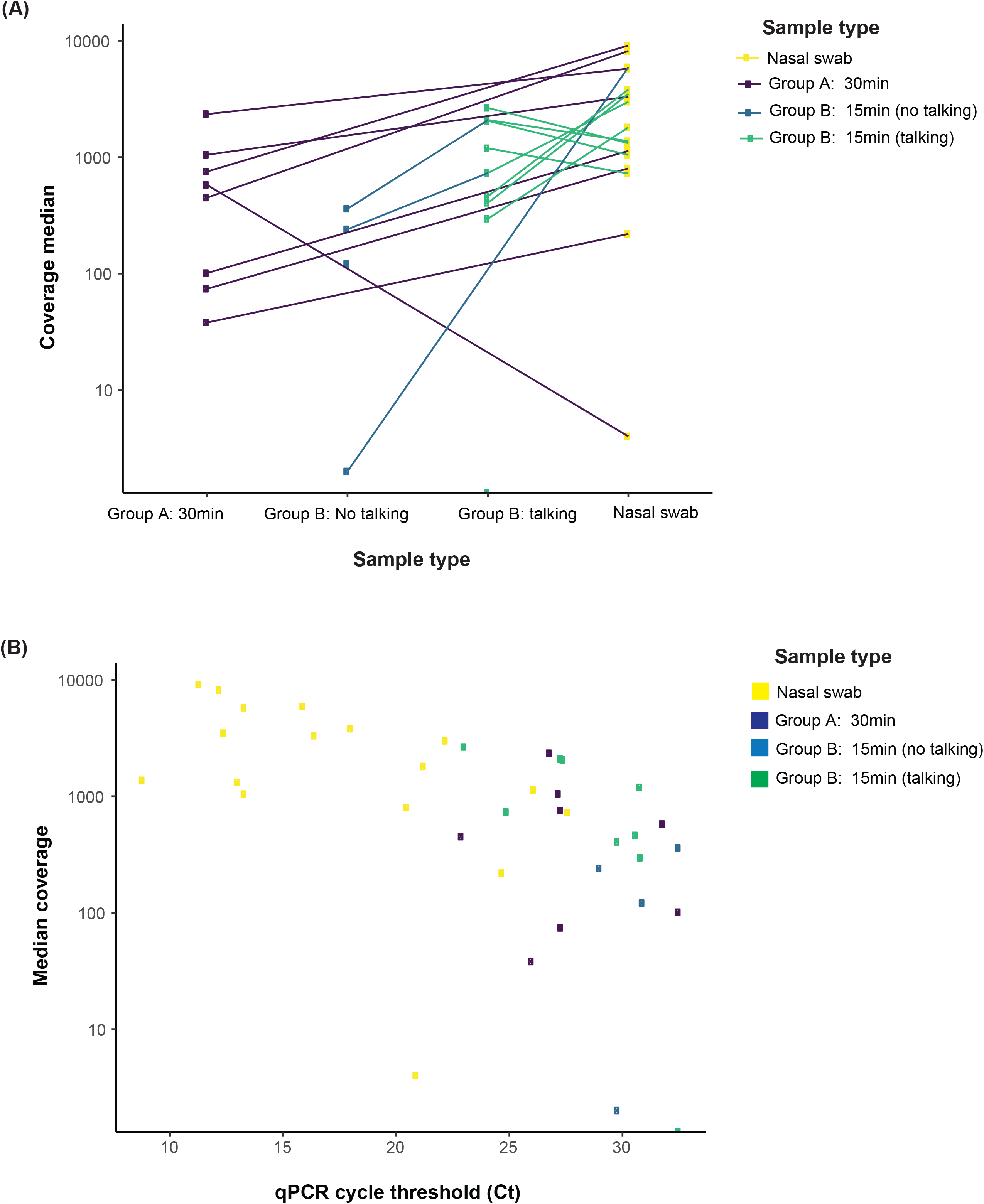
Whole genome sequencing coverage comparison between mask and swab. **(A)** The majority 71.4% (15/21) of all mask samples, including 62.5% (5/8) mask, 50.0% mask no talking, and 88.9% (8/9) mask talking samples, met our targets for sequencing coverage depth (100X) and width (>90% of the genome with >10X coverage). **(B)** Median coverage depth was correlated with viral load (r = 0.65, p < 0.001).

## Conclusions

Epidemiologic data provides strong evidence for transmission of COVID-19 through breathing or talking (12, 13). Asadi et al observed that a small proportion of individuals consistently released an order of magnitude greater aerosols than their peers (4). However, quantitative assessment of SARS-CoV-2 bioaerosol production and its determinants has been lacking. Leveraging an easy-to-use mask sampling tool, we measured interindividual variation in SARS-CoV-2 bioaerosol production and quantified the contribution of speech. We further demonstrated the potential application of bioaerosol sampling for whole genome sequencing of respiratory pathogens to study transmission and variant detection.

Overall, we observed 71% SARS-CoV-2 RNA positivity in mask samples which was significantly higher than the previously reported studies (38% to 40%) which used different sampling strategies (6,7). We found that there are orders of magnitude variation in the bioaerosol across a 15-minute period between individuals, which is significantly affected by speech and higher age. Our findings suggest that while it is challenging to define a specific threshold for risk based on cumulative exposure time (1), evaluating additional factors such as speech and physiological characteristics of the infected individual during exposure could be valuable in defining exposure risk. This was recently reflected in a report released by the CDC where they observed SARS-CoV-2 transmission in the National Football League after <15 minutes of cumulative interaction, leading to a revised definition of a high-risk contact that evaluated mask use and ventilation in addition to duration and proximity of interaction (2). The results of this study should be interpreted with the context of several limitations. RT-qPCR cannot distinguish replication-competent viruses from RNA. Our mask bioaerosol sampler captures all sizes of respiratory particles, from large to small droplets, which likely carry very different risks of infection, further dependent on proximity and ventilation. Further studies with aerodynamic particle sizers could further quantify the size distribution of SARS-CoV-2 containing bioaerosols and its determinants. Finally, to avoid discomfort among patients with higher oxygen requirements, we only recruited patients with mild symptoms.

In summary, we developed a mask sampling tool to quantify and sequence SARS-CoV-2 from exhaled breath and used our method to provide quantitative evidence on the impact of speech and interindividual variation on SARS-CoV-2 shedding in bioaerosol. We found that nasal swab viral load moderately correlates with bioaerosol production and that talking substantially amplifies exposure risk, findings which may inform assessment and risk stratification of exposures. Mask tool developed here can be further validated for its application to investigate the genomes of other respiratory pathogens.

## Data Availability

Data supporting the findings of this manuscript are available in the Supplementary Information
files or from the corresponding author upon request.

## Biographical Sketch

Dr. Verma is a molecular microbiologist at the Division of Infectious Diseases and Geographic Medicine, Stanford University School of Medicine, CA, USA. Her primary research interests include development of low-cost point-of-care diagnostics for infectious diseases.

## Acknowledgments

We thank the study teams and participants from the Covid-19 Clinical Trials Research Unit at Stanford University.

## Potential conflicts of Interest

We declare that we have no conflicts of interest for this work.

## Financial support

This study was supported by an anonymous donation to the Stanford University School of Medicine. The donors had no role in data analysis, interpretation, or decision to publish the findings.

## Figures

**Supplementary Figure 1.**
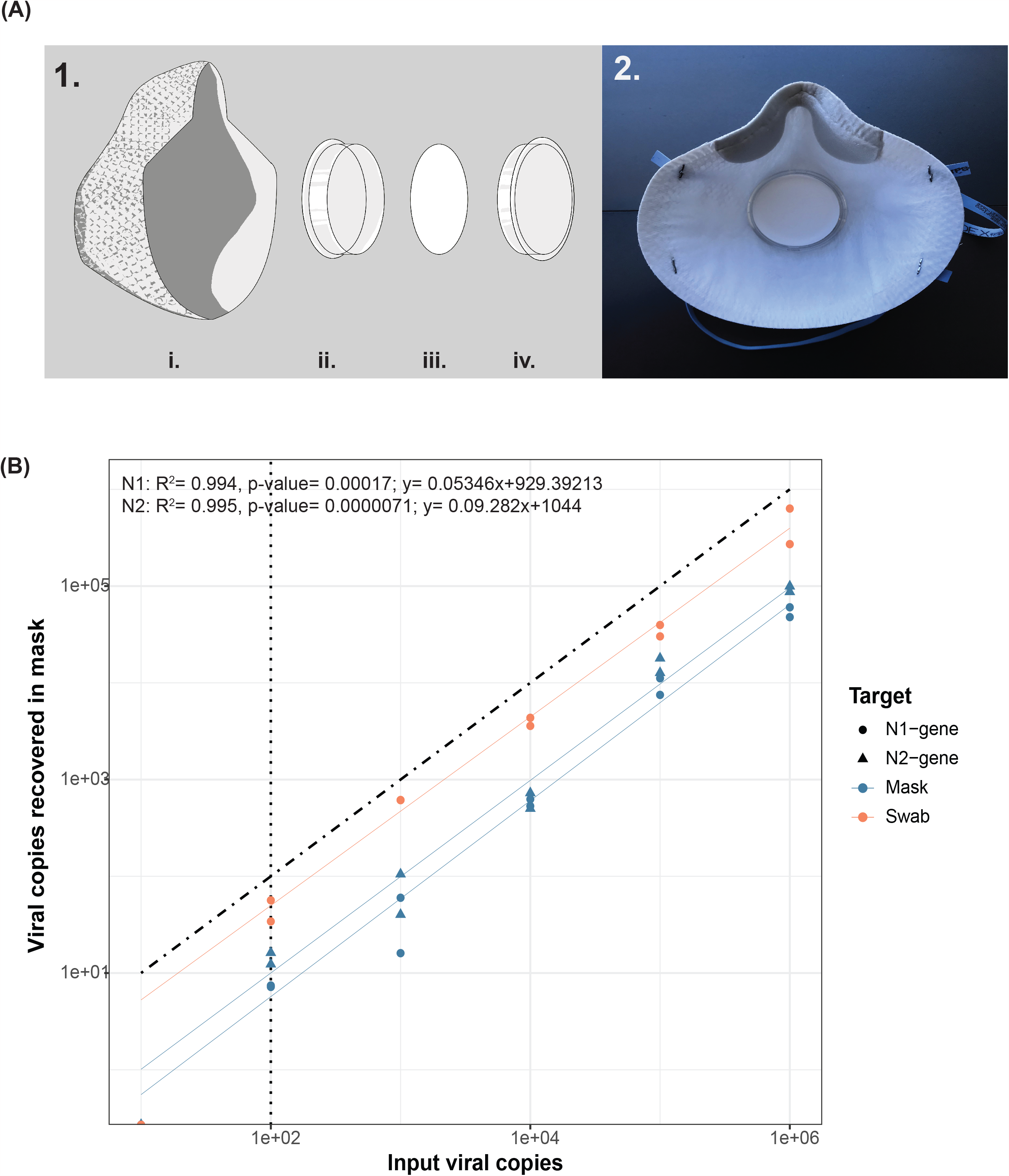
**(1)** The assembly of the gelatin filter mask. The N-95 mask (i) is used as a base, to which a 47mm petri dish (ii) is attached. A 47mm gelatin filter (iii) is placed inside, and the petri dish lid (iv) is used to secure the filter until collection. **(2)** The assembled mask. **(B)** Before exhaled breath sampling, the limit of detection (LoD) of SARS-CoV-2 RNA recovered from the gelatine filter using our sample processing protocol was compared with spiked RNA in nasal swabs. Masks and nasal swabs from healthy volunteers were spiked with serially diluted SARS CoV-2 RNA (10^6^-10^1^) in duplicates. Viral RNA was detected with as low as 100 copies/filter in both duplicates in mask and swab. Blue dots and triangles represent copies determined by the N1 and N2 assays respectively from mask. Orange dots and triangles represent copies determined by the N1 and N2 assays respectively from swab. The black, diagonal dot- dashed line represents the regression expected when the viral copies recovered are equal to the viral copies spiked. The vertical dotted line indicates the limit of detection of the assay.

## Notes

### Competing Interest Statement

The authors have declared no competing interest.

### Author Declarations

The study was approved by the Stanford IRB (#57686)

