## Supplementary material for "Variation in SARS-CoV-2 bioaerosol production in exhaled breath"

**Supplementary methods:**

**Methods:**

**Study population characteristics**

We recruited 97 COVID-19 positive patients from Stanford hospital (n=20) and Clinical Trial Research Unit (n=77) between September 2020 and March 2021. All participants were enrolled in the study within 72 hours of a positive SARS-CoV-2 RT-qPCR test. Exclusion criteria were respiratory rate >20 breaths per minute, room air oxygen saturation <94%, pregnancy or breastfeeding. The exhaled breath samples were collected on the first day of enrollment. Median age of the participants was 44.5 years (IQR, 36-57), and 46.3% (45/97) were female. Symptom data was available for 91 patients and the majority of participants (88/91; 96.7%) reported one or more COVID-19 related symptoms several days prior to enrollment (median, 4 days; range, 0-13). Symptoms with onset more than three weeks prior to study enrollment were not considered to be associated with COVID-19.

**Mask design and analytical validation**

We fitted N95 masks (Moldex, 220 NSN series) with a 47mm Petri dish (Fisherbrand™ Sterile Polystyrene Petri Dish) containing dissolvable gelatin membrane filter disc (Sartorius, Germany) (**Supplementary figure 1**). To assess the stability and recovery of viral RNA from the filter, we spiked serially diluted (1x10^6^-1x10^1^ copies) synthetic SARS-CoV-2 RNA (Twist Bioscience, San Francisco, CA) on the mask and compared with spiked nasal swabs samples from healthy volunteers. All spiking experiments were performed in duplicates. The filter was directly dissolved on the Petri dish by adding 1ml of Primestore MTM (Longhorn Vaccines & Diagnostics) viral inactivation, RNA stabilizing media. RNA was extracted using MagMAX™ Viral/Pathogen Ultra Kit (Cat # A42356 Applied Biosystems). RNA was eluted in 50ul nuclease-free water.

**Exhaled breath sample collection**

We collected 141 mask samples from 97 individuals recruited in two groups (A & B) with RT-PCR-confirmed SARS-CoV-2 infection. Group A (n=53) was instructed to wear a mask for 30 minutes and was allowed to talk (or not) without further prompting. Group B (n=44) was instructed to wear two masks for 15 minutes each. For the first mask, they were instructed not to talk, while for the second mask they were asked to talk with the interviewer or a family member. Additionally, we enrolled 22 healthy volunteers under group A (n=9) and B (n=13) respectively. After use, the filter was removed and analyzed as described above.

**RT-PCR assay for SARS-CoV-2 RNA**

We performed RT-qPCR using the CDC qualified primers and probes amplifying N1 and N2 regions of SARS-CoV-2 N-gene and human RNaseP as a quality control (1). TaqPath one-step RT-PCR mastermix (Invitrogen, Darmstadt, Germany) was used in a 20ul reaction volume and the samples were analyzed on a StepOne-Plus (Applied Biosystems) instrument, using the following program: 10 min at 50 °C for reverse transcription, followed by 3 min at 95 °C and 40 cycles of 10 s at 95 °C, 15 s at 56 °C, and 5 s at 72 °C. We estimated copies/sample from a standard curve using a pET21b+ plasmid (GenScript, USA) with the N-gene. The cycle threshold (Ct) cutoff for positive samples was <38.

**Whole genome sequencing of SARS-CoV-2 from exhaled breath**

We sequenced SARS-CoV-2 genome from 21 (9 talking, 4 no talking and 8 without any maneuver) exhaled breath samples (median Ct= 29; range 22.8-32.5) and 14 paired nasal swab samples (median Ct= 8.2; range 8.8-26.1). We adapted ARTIC V3 Illumina sequencing protocol described previously to sequence SARS-CoV-2 (2). The multiplex PCR primers set v3, separated into two pools (A and B), was used to span the whole genome (https://artic.network/ncov-2019). Briefly, 10μL template RNA was converted to cDNA using Lunascript RT supermix (New England Biolabs, Ipswich, MA). cDNA was amplified using each of the two ARTIC v3 primer pools, 1X Q5 Hotstart mastermix (New England Biolabs, Ipswich, MA). The purified, pooled amplicons were dAtail repaired with NEBNext Ultra II End prep enzyme mix. Adapter ligation was performed using with NEBNext Ultra II ligation mix and 15uM NEBNext Adaptor. Dual indexing was performed using i5 and i7 adapters (NEB) as per the manufacturer’s instructions using KAPA Hifi hotstart mastermix. The pooled samples were analyzed on Illumina MiSeq using V2 reagent kit 500cycles (212/212 cycles).

**Whole genome sequencing analysis pipeline**

We used the nfcore/viralrecon bioinformatic pipeline containerized on Nextflow to perform variant calling and generate consensus sequences from raw reads (3). Briefly, we used this pipeline to remove reads mapping to the host genome with Kraken2 (4) align reads to the MN908947.3 reference genome with Bowtie 2 (5), remove primer sequences with iVar (6) call variants with respect to the reference genome with iVar, generate a consensus sequence with iVar, and assign Pango lineage with pangolin (7). We aligned consensus genomes with mafft (8) and masked previously reported problematic sites (9). We used the R package ape to measure pairwise SNP distance between consensus sequences (10).

**Statistical analysis**

In paired masks (talking, not talking) from the same individual, we compared SARS-CoV-2 detection dichotomously using McNemar’s test and quantitatively using Wilcoxon sign rank tests. We calculated Pearson’s correlation coefficient (r) comparing viral copies estimated from nasal swab and mask aerosol samples.
